# Outcomes of Percutaneous Coronary Intervention for Protected versus Unprotected Left Main Coronary Artery Disease: Insights from the VA CART Program

**DOI:** 10.1101/2023.10.27.23297698

**Authors:** Pedro Engel Gonzalez, Annika Hebbe, Yasin Hussain, Rohan Khera, Subhash Banerjee, Mary E Plomondon, Stephen W. Waldo, Steven E. Pfau, Jeptha P. Curtis, Samit M. Shah

**Author notes:** **Sources of Support:** Dr. Khera received support from the National Heart, Lung, and Blood Institute of the National Institutes of Health under the award K23HL153775-01A1. The funders had no role in the design and conduct of the study; collection, management, analysis, and interpretation of the data; preparation, review, or approval of the manuscript; and decision to submit the manuscript for publication. **Conflicts of Interest:** Dr. Shah receives investigator-initiated research support from Abbott Vascular and the Food and Drug Administration. Dr. Waldo has received investigator-initiated research support from Abiomed, Cardiovascular Systems Incorporated, Janssen Pharmaceuticals, National Institutes of Health and VA Health Services Research & Development. Dr. Curtis receives salary support from the American College of Cardiology for his role as Chief Scientific Advisor of the National Cardiovascular Data Registry. Dr Curtis has equity interest in Medtronic, a maker of cardiac stents. **Corresponding author:** Samit Shah, MD PhD, 950 Campbell Avenue, West Haven, CT 06516.

## Abstract

**Background:** Practice patterns and outcomes of protected left main (PLM) and unprotected left main (ULM) percutaneous coronary intervention (PCI), as well as the differences between these types of PCI, are not well defined in real-world clinical practice.

**Methods:** Data collected from all Veteran Affairs (VA) catheterization laboratories participating in the Clinical Assessment Reporting and Tracking Program between 2009 and 2019. The analysis included 4,351 patients undergoing left main PCI, of which 1,306 pairs of PLM and ULM PCI were included in a propensity matched cohort. Patients and procedural characteristics were compared between PLM and ULM PCI. Temporal trends were also assessed. Peri-procedural and one-year major adverse cardiovascular events (MACE) were compared using cumulative incidence plots. The primary outcome was MACE outcomes at 1-year, which was defined as a composite of all-cause mortality, rehospitalization for myocardial infarction (MI), rehospitalization for stroke or urgent revascularization.

**Results:** ULM PCI patients in comparison to PLM PCI were older (71.5 vs 69.2; P < 0.001), more clinically complex and more likely to present with ACS. In the propensity matched cohort, radial access was used more often for ULM PCI (21% [273] vs. 14% [185], P < 0.001), and ULM PCI was more likely to involve the LM bifurcation (22% vs 14%; P = 0.003) and require mechanical circulatory support (10% [134] vs 1% [17]; P <0.001). One-year MACE occurred more frequently with ULM PCI compared to PLM PCI (22% [289] vs. 16% [215]; P = < 0.001) and all-cause mortality was also higher (16% [213] vs. 10% [125]; P = < 0.001). In the matched cohort there was a low incidence of rehospitalization for MI (4% [48] ULM vs. 4% [48] PLM; P = 1.000) or revascularization (7% [94] ULM vs. 6% [84] PLM; P = 0.485).

**Conclusions:** Veterans undergoing PLM PCI had better one-year outcomes than those undergoing ULM PCI, but in both groups there was a high rate of mortality and MACE at one-year despite a relatively low rate of MI or revascularization.

---

- What is new?

- In a large multi-center sample from the United States Department of Veterans Affairs, there has been increasing use of left main PCI in patients who have not undergone prior coronary artery bypass graft surgery (CABG).
- Compared to patients with prior CABG who underwent left main PCI, those who underwent left main PCI without prior CABG had a higher rate of major adverse cardiovascular events and all cause death at 12 months.
- While there was an overall low rate of rehospitalization for myocardial infarction or revascularization, there was a higher 12-month incidence of major adverse cardiovascular events in this real-world population compared to clinical trial populations.
- What are the clinical implications?

- Veterans who underwent left main PCI in real-world practice were more medically complex and higher risk than those who were enrolled in clinical trials of left main PCI
- Patients who undergo left main PCI may have been ineligible for coronary artery bypass graft surgery (“surgical turn down”, and thus may represent a higher risk population
- In elective cases, shared decision-making regarding the best method of coronary revascularization should include a discussion of the patient’s 1-year prognosis

## Introduction

Severe left main (LM) coronary artery disease (CAD) is found in approximately 4% of all patients undergoing coronary angiography.^1,2^ Coronary artery bypass graft (CABG) surgery has long been the standard of care for patients with LM CAD, whereas percutaneous coronary intervention (PCI) has historically been performed only for patients at prohibitive or high-risk for surgical intervention.^3^ However, recent randomized clinical trials suggest that PCI for LM CAD may be an effective and safe alternative to CABG.^4–7^ LM CAD is classified based on the presence or absence of a prior bypass graft. Protected left main (PLM) PCI is defined as percutaneous revascularization for LM CAD in post-CABG patients with a patent bypass graft to a branch vessel arising from the left main (usually the left anterior descending artery, left circumflex/obtuse marginal, or ramus intermedius), whereas unprotected left main (ULM) PCI refers to treatment of the LM in the absence of prior bypass graft to the left coronary system. A recent analysis from the National Cardiovascular Data Registries (NCDR) Cath PCI registry showed that ULM PCI represented only a small proportion of all PCIs (1.0%) and the in-hospital rates of mortality, MI, stroke, and emergent CABG were significantly higher in comparison to “all other” non-LM PCI (26% vs. 9%; P < 0.001).^8^

PLM PCI has not been well studied in clinical trials and there has not been a dedicated contemporary evaluation of outcomes despite PLM PCI being performed in clinical practice. In the era of bare metal stents, the one-year major adverse cardiovascular event (MACE) rate for PLM PCI was 25-29%^9,10,11^ Studies with first generation drug eluting stents have shown better outcomes for PLM PCI compared with ULM PCI^10,12^; however, the German Cypher Stent registry showed no significant differences in the incidence of MACE at 6-months (the primary end-point occurring in 13-14% of patients).^13^

To better characterize utilization and real-world outcomes of ULM and PLM PCI in the Veterans administration, we studied patient and procedural characteristics associated with ULM and PLM PCI in the Veteran Affairs (VA) Clinical Assessment Reporting and Tracking (CART) Program between 2009 and 2019, as well as the one-year clinical outcomes of patients undergoing both types of procedures.

## Methods

### Cohort

The cohort was derived from the VA CART Program, a national quality and safety program supporting all VA cardiac catheterization laboratories. The CART Program uses a software application embedded into the electronic health record to collect standardized data on all invasive cardiac procedures performed at VA facilities across the United States, including coronary angiography and PCI.^14^ Data from coronary angiograms and/or PCI are entered at each institution using a standardized interface. Quality checks of CART data are periodically conducted for completeness and accuracy. The CART data are also merged with the VA Computerized Patient Record System (CPRS) electronic health record to derive longitudinal patient data.^15^ Institutional review board approval was obtained by the Colorado Multiple Institutions Review Board and with a waiver of informed consent.

We identified all PCI of the left main procedures performed at VA catheterization laboratories from January 1^st^, 2009 to September 30^th^, 2019. PLM PCI was defined as any PCI of the LM in patients with history of prior CABG and presence of at least one functioning arterial or venous graft supplying the left coronary circulation, whereas ULM PCI was defined as any PCI of the LM in patients with no history of left-sided grafts providing protection to the LM or documented occlusion of prior left-sided graft(s). Procedures were excluded if patients were under the age of 18 or underwent balloon angioplasty alone, which may have been performed as a temporizing measure prior to surgical revascularization.

### Covariates of Interest

Baseline patient demographic, clinical, and procedural characteristics were collected from the registry and compared between patients undergoing PLM PCI and ULM PCI. Temporal trends of ULM and PLM PCI over time were assessed both by total patient count and percentage of all LM PCI. Patient characteristics and comorbidities associated with outcomes were selected.

Procedural and anatomic data was also collected. The estimated patient clinical complexity was derived from pre-procedure clinical factors and calculated using models developed by the NCDR Cath PCI simplified risk score, which describes the risk of early mortality following PCI.^16^ The “VA Syntax Score” simplified anatomic complexity scores were automatically derived based on data provided by individual operators interpreting each angiogram utilizing methods previously described, which has a moderate correlation with a manually computed SYNTAX score and is associated with an increased hazard of adverse events among patients undergoing PCI.^17^ Of note, clinical outcomes related to the anatomic complexity score have been validated for individuals with native vessel disease only. Institutional annual PCI volumes (including LM and non-LM PCI) were also obtained and tertiles were created to categorize institutions based on volume.

### Outcomes

The primary outcome was MACE outcomes at one-year, which was defined as a composite of all-cause mortality, rehospitalization for myocardial infarction (MI), rehospitalization for stroke, or urgent revascularization. The primary outcome, and its individual components, were obtained via a merged examination of the VA CART data and CPRS (the VA electronic health record) to derive longitudinal patient data.^14^ One year of follow-up data was available for all subjects in the cohort.

Peri-procedural outcomes were also collected from the VA CART data, including periprocedural MACE which was defined as all-cause mortality, MI, stroke, or emergent CABG.

### Statistical Analysis

Patient, procedural, and institutional characteristics for patients undergoing PLM and ULM PCI were compared using *t* tests for continuous variables and *χ*^2^ or Fisher exact tests for categorical values. In the assessment of outcomes across the two groups, to reduce potential confounding due to differences in patient characteristics, we performed one-to-one propensity score matching by a greedy matching algorithm with a caliper of 0.1 was performed based on the following variables: age, gender, race, body mass index (BMI), hemoglobin, left ventricular ejection fraction (LVEF), prior MI, prior PCI, diabetes mellitus, peripheral vascular disease, renal dysfunction, New York Heart Association (NYHA) class, procedure status, chronic lung disease, glomerular filtration rate (GFR), hypertension, tobacco use, family history of coronary artery disease, presentation of ST-segment elevation myocardial infarction (STEMI), non-ST-segment elevation myocardial infarction (NSTEMI), unstable angina, and stable angina. Balance between the groups of the matched cohort were assessed by calculating standardized differences for which a difference of less than 0.10 was considered to indicate good balance.

One-year outcome analyses were conducted for MACE, all-cause mortality, rehospitalization for MI and rehospitalization for revascularization within the matched cohort. Cumulative incidence plots were derived for all outcomes. Cox proportional hazard models with robust variance estimators to account for matched nature of the cohort were fit for MACE and all-cause mortality. To account for the competing risk of mortality, competing risk analyses were performed by fitting both a clustered Fine-Gray model and a cause-specific hazard model with a robust variance estimator for MI and revascularization. A sensitivity analysis was performed to determine the effects of an unmeasured confounders and reported as an E-value which represents the minimum strength of an unmeasured confounder in relation to the treatment and outcome to explain away the observed association between the treatment and outcome.. If the strength of unmeasured confounding is less than the calculated E-value, then the association between the treatment and outcome could not be overturned by the unmeasured confounder. There is no specific guidance on what is considered a small or large E-value, and that magnitude should be interpreted in the context of this paper only. ^18^

## Results

### Cohort

A total of 4,584 patients underwent left main PCI between 2009 and 2019 at VA medical centers. Of these patients, 233 were excluded for undergoing balloon angioplasty alone without stenting. Of the remaining 4,351 LM PCI, 2,783 were PLM PCI and 1,568 were ULM PCI, of which 1,306 PLM and ULM PCI were included in the propensity matched cohort (**Figure 1**). All variables included in the propensity score matching resulted in standardized mean difference < 0.10, indicating good balance and therefore no further adjustments were made in the models (**Supplemental Table S1)**.

**Figure 1.**
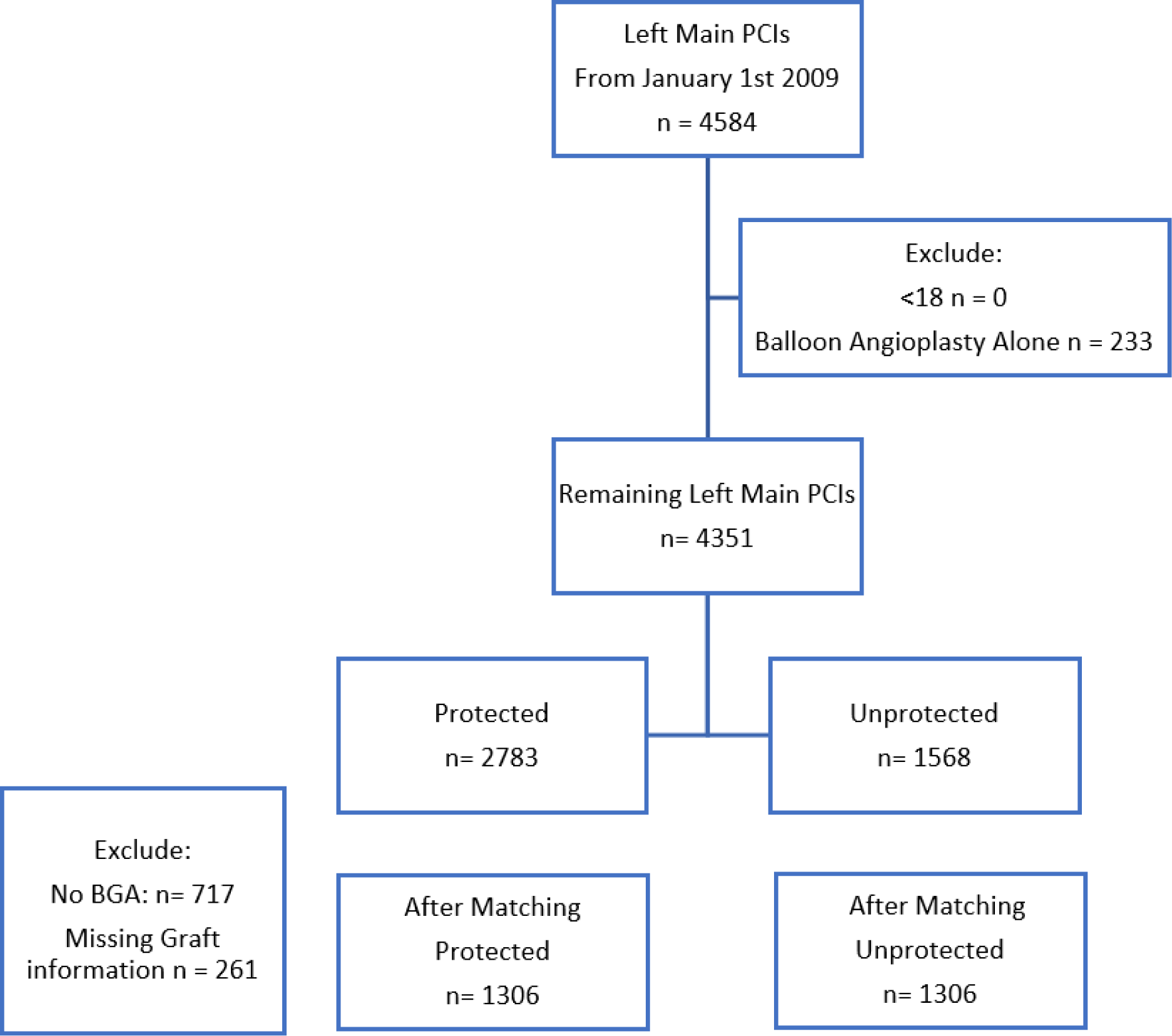
Study flow diagram. The flow diagram detailing the results of the screening process of identifying PLM and ULM PCI cases, and the creation of a one-to-one propensity score matched cohort. PCI = percutaneous coronary intervention; PLM = Protected Left Main left main; ULM = Unprotected left main; BGA = bypass graft angiography.

### Demographic, Clinical, Procedural, and Institutional Characteristics

In the unmatched cohort at baseline, patients undergoing ULM PCI had notable differences in comparison to PLM PCI patients (**Table 1**). The ULM PCI patients were older (71.5 vs 69.2; P < 0.001), had higher burden of chronic lung disease (41% [650] vs. 33% [593]; P <0.001), renal dysfunction (9% [138] vs. 4% [78]; P < 0.001), more advanced NYHA class (between-group difference P <0.001), were more likely to present for urgent (33% [516] vs. 30% [546]) or emergent/salvage PCI (11% [178] vs. 4% [64]), more likely to present as a NSTEMI (27% [428] vs. 18% [333]; P <0.001) or STEMI (6% [87] vs. 2% [41]; P <0.001), and had a higher NCDR clinical complexity score (26.6 vs. 21.1; P < 0.001). On the other hand, patients in the ULM PCI group were less likely to have comorbidities such as prior MI (46% [716] vs. 62% [1123]; P < 0.001), and diabetes 49% [770] vs. 61% [1095]; P <0.001).

**Table 1.**
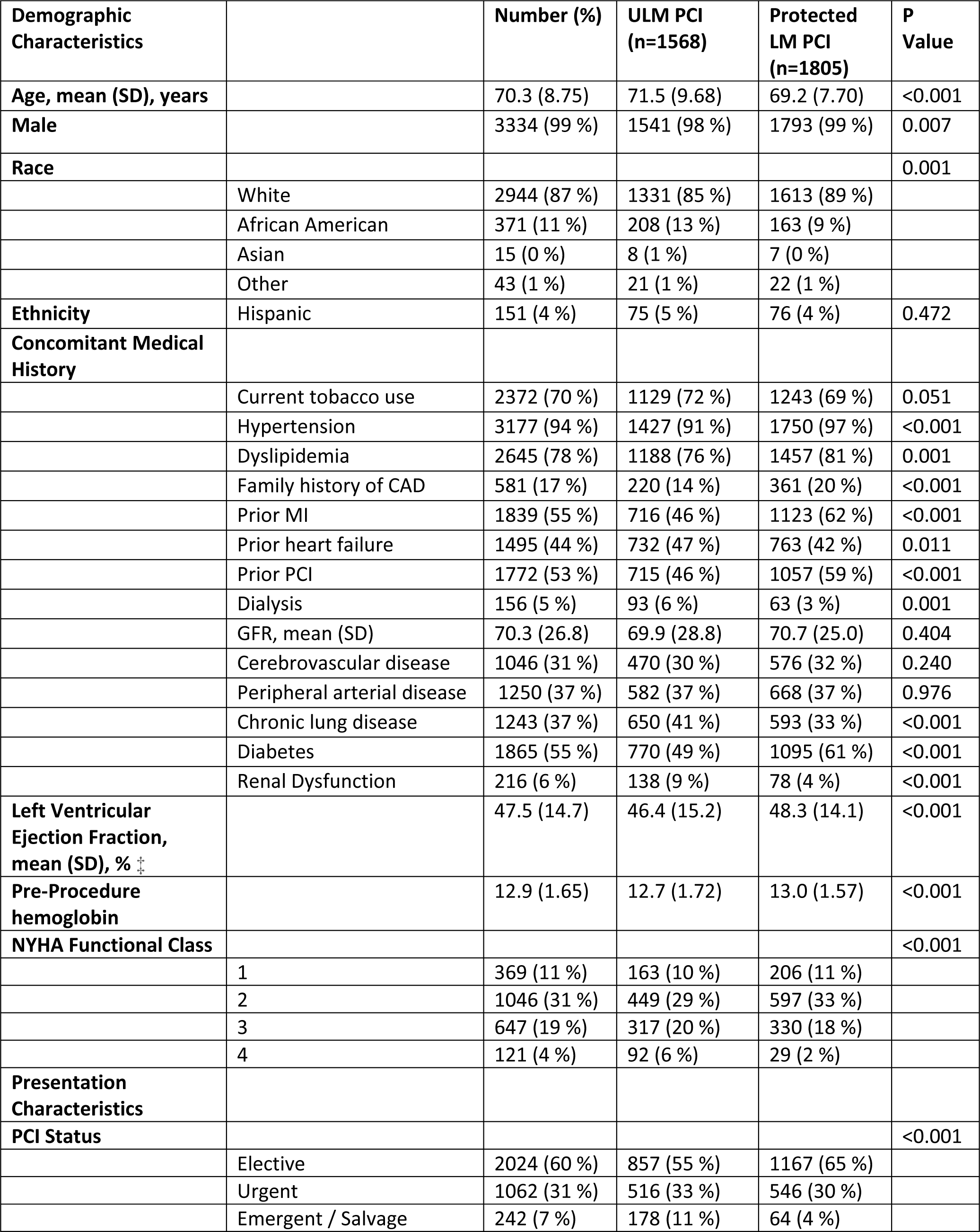
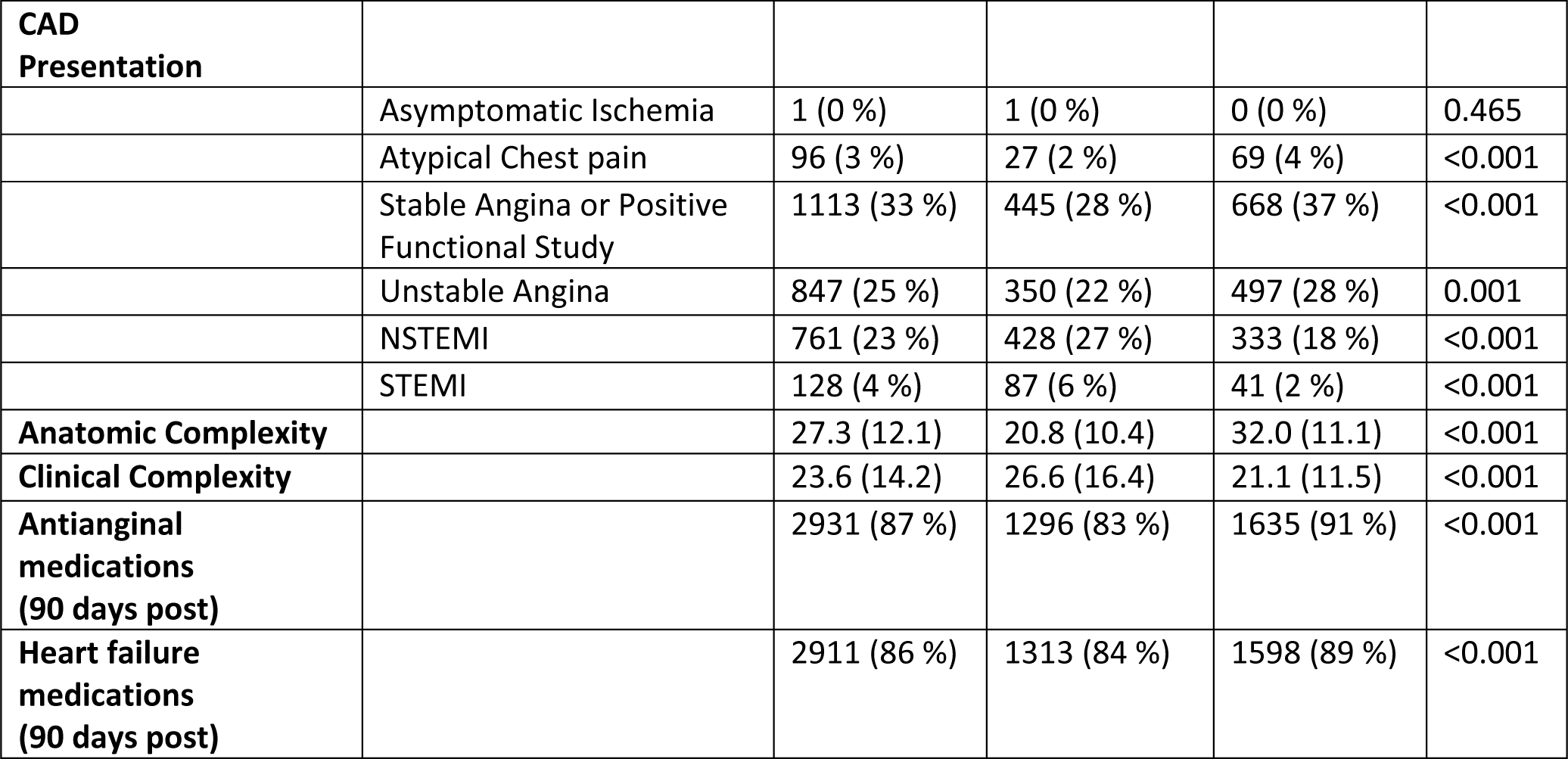
Patient Demographic and Clinical Characteristics for the Unmatched Cohort. Values are mean ± standard deviation (SD) or n (%). CAD = coronary artery disease; NYHA = New York Heart Association; GFR = glomerular filtration rate; NSTEMI = non-ST-segment elevation myocardial infarction; STEMI = ST-segment elevation myocardial infarction

| Demographic Characteristics |  | Number (%) | ULM PCI (n=1568) | Protected LM PCI (n=1805) | P Value |
| --- | --- | --- | --- | --- | --- |
| Age, mean (SD), years |  | 70.3 (8.75) | 71.5 (9.68) | 69.2 (7.70) | <0.001 |
| Male |  | 3334 (99 %) | 1541 (98 %) | 1793 (99 %) | 0.007 |
| Race |  |  |  |  | 0.001 |
|  | White | 2944 (87 %) | 1331 (85 %) | 1613 (89 %) |  |
|  | African American | 371 (11 %) | 208 (13 %) | 163 (9 %) |  |
|  | Asian | 15 (0 %) | 8 (1 %) | 7 (0 %) |  |
|  | Other | 43 (1 %) | 21 (1 %) | 22 (1 %) |  |
| Ethnicity | Hispanic | 151 (4 %) | 75 (5 %) | 76 (4 %) | 0.472 |
| Concomitant Medical History |  |  |  |  |  |
|  | Current tobacco use | 2372 (70 %) | 1129 (72 %) | 1243 (69 %) | 0.051 |
|  | Hypertension | 3177 (94 %) | 1427 (91 %) | 1750 (97 %) | <0.001 |
|  | Dyslipidemia | 2645 (78 %) | 1188 (76 %) | 1457 (81 %) | 0.001 |
|  | Family history of CAD | 581 (17 %) | 220 (14 %) | 361 (20 %) | <0.001 |
|  | Prior MI | 1839 (55 %) | 716 (46 %) | 1123 (62 %) | <0.001 |
|  | Prior heart failure | 1495 (44 %) | 732 (47 %) | 763 (42 %) | 0.011 |
|  | Prior PCI | 1772 (53 %) | 715 (46 %) | 1057 (59 %) | <0.001 |
|  | Dialysis | 156 (5 %) | 93 (6 %) | 63 (3 %) | 0.001 |
|  | GFR, mean (SD) | 70.3 (26.8) | 69.9 (28.8) | 70.7 (25.0) | 0.404 |
|  | Cerebrovascular disease | 1046 (31 %) | 470 (30 %) | 576 (32 %) | 0.240 |
|  | Peripheral arterial disease | 1250 (37 %) | 582 (37 %) | 668 (37 %) | 0.976 |
|  | Chronic lung disease | 1243 (37 %) | 650 (41 %) | 593 (33 %) | <0.001 |
|  | Diabetes | 1865 (55 %) | 770 (49 %) | 1095 (61 %) | <0.001 |
|  | Renal Dysfunction | 216 (6 %) | 138 (9 %) | 78 (4 %) | <0.001 |
| Left Ventricular Ejection Fraction, mean (SD), % ‡ |  | 47.5 (14.7) | 46.4 (15.2) | 48.3 (14.1) | <0.001 |
| Pre-Procedure hemoglobin |  | 12.9 (1.65) | 12.7 (1.72) | 13.0 (1.57) | <0.001 |
| NYHA Functional Class |  |  |  |  | <0.001 |
|  | 1 | 369 (11 %) | 163 (10 %) | 206 (11 %) |  |
|  | 2 | 1046 (31 %) | 449 (29 %) | 597 (33 %) |  |
|  | 3 | 647 (19 %) | 317 (20 %) | 330 (18 %) |  |
|  | 4 | 121 (4 %) | 92 (6 %) | 29 (2 %) |  |
| Presentation Characteristics |  |  |  |  |  |
| PCI Status |  |  |  |  | <0.001 |
|  | Elective | 2024 (60 %) | 857 (55 %) | 1167 (65 %) |  |
|  | Urgent | 1062 (31 %) | 516 (33 %) | 546 (30 %) |  |
|  | Emergent / Salvage | 242 (7 %) | 178 (11 %) | 64 (4 %) |  |

**Table 1. Patient Demographic and Clinical Characteristics for the Unmatched Cohort.**
| **CAD Presentation** |  |  |  |  |  |
| --- | --- | --- | --- | --- | --- |
|  | Asymptomatic Ischemia | 1 (0 %) | 1 (0 %) | 0 (0 %) | 0.465 |
|  | Atypical Chest pain | 96 (3 %) | 27 (2 %) | 69 (4 %) | <0.001 |
|  | Stable Angina or Positive Functional Study | 1113 (33 %) | 445 (28 %) | 668 (37 %) | <0.001 |
|  | Unstable Angina | 847 (25 %) | 350 (22 %) | 497 (28 %) | 0.001 |
|  | NSTEMI | 761 (23 %) | 428 (27 %) | 333 (18 %) | <0.001 |
|  | STEMI | 128 (4 %) | 87 (6 %) | 41 (2 %) | <0.001 |
| **Anatomic Complexity** |  | 27.3 (12.1) | 20.8 (10.4) | 32.0 (11.1) | <0.001 |
| **Clinical Complexity** |  | 23.6 (14.2) | 26.6 (16.4) | 21.1 (11.5) | <0.001 |
| **Antianginal medications (90 days post)** |  | 2931 (87 %) | 1296 (83 %) | 1635 (91 %) | <0.001 |
| **Heart failure medications (90 days post)** |  | 2911 (86 %) | 1313 (84 %) | 1598 (89 %) | <0.001 |

In the propensity matched cohort comparing patients undergoing ULM and PLM PCI, there were no significant differences in terms of age (70.0 vs 69.5; P = 0.151), sex (male 99% [1,296] vs. 99% [1,297]; P = 1.000), and race/ethnicity (**Table 2**). There was a high burden of medical comorbidities in both groups, including current tobacco use (71% [923] vs. 70% [914]; P = 0.731), prior MI (55% [716] vs. 60% [780]; P = 0.0.088), prior heart failure (45% [582] vs. 43% [561]; P = 0.430), diabetes (56% [733] vs. 58% [762]; P = 0.269), peripheral arterial disease (37% [482] vs. 37% [480]; P = 0.968), and chronic lung disease (36% [467] vs. 34% [443]; P = 0.0.345). The LVEF was also similar in both groups (47.4% vs 48.1%; P = 0.240) and there was no significant difference in NYHA class (P = 0.826). There were no between group differences in PCI status (P = 0.333); elective (62% [805] vs. 64% [831]), urgent (32% [412] vs. 31% [404]), emergent/salvage (6% [72] vs. 4% [53]). There were no significant differences in the proportion of ACS presentation; STEMI (3% [36] vs. 3% [33]; P = 0.807), NSTEMI (22% [285] vs. 20% [256]; P = 0.176), or unstable angina (34% [445] vs. 35% [460]; P = 0.593). Patients undergoing ULM PCI had a higher NCDR clinical complexity risk score (22.9 vs 21.6; P < 0.001).

**Table 2.** Patient Demographic and Clinical Characteristics for the Matched Cohort. Values are mean ± standard deviation (SD) or n (%) Abbreviations as per Table 1.

| Demographic Characteristics |  | Number (%) | ULM PCI (n=1306) | Protected LM PCI (n=1306) | P Value | SMD |
| --- | --- | --- | --- | --- | --- | --- |
| Age, mean (SD), years |  | 69.7 (8.34) | 70.0 (8.90) | 69.5 (7.74) | 0.151 | 0.056 |
| Male |  | 2593 (99 %) | 1296 (99 %) | 1297 (99 %) | 1.000 | 0.009 |
| Race |  |  |  |  | 0.772 |  |
|  | White | 2306 (88 %) | 1145 (88 %) | 1161 (89 %) |  | 0.038 |
|  | African American | 263 (10 %) | 138 (11 %) | 125 (10 %) |  | 0.022 |
|  | Asian | 12 (0 %) | 7 (1 %) | 5 (0 %) |  | 0.023 |
|  | Other | 31 (1 %) | 16 (1 %) | 15 (1 %) |  | 0.007 |
| Ethnicity | Hispanic | 112 (4 %) | 58 (4 %) | 54 (4 %) | 0.772 | 0.015 |
| Concomitant Medical History |  |  |  |  |  |  |
|  | Current tobacco use | 1837 (70 %) | 923 (71 %) | 914 (70 %) | 0.731 | 0.015 |
|  | Hypertension | 2513 (96 %) | 1249 (96 %) | 1264 (97 %) | 0.151 | 0.060 |
|  | Dyslipidemia | 2088 (80 %) | 1030 (79 %) | 1058 (81 %) | 0.187 | 0.054 |
|  | Family history of CAD | 462 (18 %) | 220 (17 %) | 242 (19 %) | 0.611 | 0.022 |
|  | Prior MI | 1496 (57 %) | 716 (55 %) | 780 (60 %) | 0.088 | 0.068 |
|  | Prior heart failure | 1143 (44 %) | 582 (45 %) | 561 (43 %) | 0.430 | 0.032 |
|  | Prior PCI | 1444 (55 %) | 701 (54 %) | 743 (57 %) | 0.107 | 0.065 |
|  | Dialysis | 110 (4 %) | 56 (4 %) | 54 (4 %) | 0.922 | 0.008 |
|  | GFR, mean (SD) ‡ | 70.4 (25.7) | 70.6 (25.9) | 70.2 (25.5) | 0.728 | 0.014 |
|  | Cerebrovascular disease | 823 (32 %) | 402 (31 %) | 421 (32 %) | 0.448 | 0.031 |
|  | Peripheral arterial disease | 962 (37 %) | 482 (37 %) | 480 (37 %) | 0.968 | 0.003 |
|  | Chronic lung disease | 910 (35 %) | 467 (36 %) | 443 (34 %) | 0.345 | 0.039 |
|  | Diabetes | 1495 (57 %) | 733 (56 %) | 762 (58 %) | 0.269 | 0.045 |
|  | Renal Dysfunction | 137 (5 %) | 70 (5 %) | 67 (5 %) | 0.936 | 0.006 |
| Left Ventricular Ejection Fraction, mean (SD), % |  | 47.7 (14.5) | 47.4 (14.9) | 48.1 (14.1) | 0.240 | 0.046 |
| Pre-Procedure hemoglobin |  | 12.9 (1.62) | 12.9 (1.65) | 12.9 (1.58) | 0.488 | 0.027 |
| NYHA Functional Class |  |  |  |  | 0.826 |  |
|  | 1 | 286 (11 %) | 143 (11 %) | 143 (11 %) |  | <0.001 |
|  | 2 | 841 (32 %) | 415 (32 %) | 426 (33 %) |  | 0.018 |
|  | 3 | 494 (19 %) | 250 (19 %) | 244 (19 %) |  | 0.012 |
|  | 4 | 50 (2 %) | 29 (2 %) | 21 (2 %) |  | 0.045 |
| **Presentation Characteristics** |  |  |  |  |  |  |
| **PCI Status** |  |  |  |  | 0.333 |  |
|  | Elective | 1636 (63 %) | 805 (62 %) | 831 (64 %) |  | 0.041 |
|  | Urgent | 816 (31 %) | 412 (32 %) | 404 (31 %) |  | 0.013 |
|  | Emergent / Salvage | 125 (5 %) | 72 (6 %) | 53 (4 %) |  | 0.068 |
| **CAD Presentation** |  |  |  |  |  |  |
|  | Asymptomatic Ischemia | 1 (0 %) | 1 (0 %) | 0 (0 %) | 1.000 | 0.039 |
|  | Atypical Chest pain | 73 (3 %) | 27 (2 %) | 46 (4 %) | 0.081 | 0.073 |
|  | Stable Angina or Positive Functional Study | 905 (35 %) | 445 (34 %) | 460 (35 %) | 0.593 | 0.023 |
|  | Unstable Angina | 699 (27 %) | 340 (26 %) | 359 (27 %) | 0.426 | 0.033 |
|  | NSTEMI | 541 (21 %) | 285 (22 %) | 256 (20 %) | 0.176 | 0.055 |
|  | STEMI | 69 (3 %) | 36 (3 %) | 33 (3 %) | 0.807 | 0.014 |
| **Anatomic Complexity** |  | 28.2 (12.2) | 24.3 (11.9) | 31.7 (11.4) | 0.007 | 0.099 |
| **Clinical Complexity** |  | 22.3 (12.8) | 22.9 (13.5) | 21.6 (12.0) | <0.001 | 0.636 |
| **Cardiogenic Shock** |  | 0 (0 %) | 0 (0 %) | 0 (0 %) | - | <0.001 |
| **Arrhythmia** |  | 5 (0 %) | 4 (0 %) | 1 (0 %) | 0.375 | 0.053 |
| **Heart Failure** |  | 4 (0 %) | 2 (0 %) | 2 (0 %) | 1.000 | <0.001 |
| **Cardiomyopathy** |  | 30 (1 %) | 25 (2 %) | 5 (0 %) | <0.001 | 0.144 |
| **Aortic Valve Disease** |  | 2 (0 %) | 1 (0 %) | 1 (0 %) | 1.000 | <0.001 |
| **Mitral Valve Disease** |  | 1 (0 %) | 0 (0 %) | 1 (0 %) | 1.000 | 0.039 |
| **Antianginal medications (90 days post)** |  | 2314 (89 %) | 1129 (86 %) | 1185 (91 %) | 0.001 | 0.135 |
| **Heart failure medications (90 days post)** |  | 2292 (88 %) | 1133 (87 %) | 1159 (89 %) | 0.136 | 0.061 |

There were significant between group differences regarding arterial access site with more frequent use of radial access (21% [273] vs. 14% [185]) compared to femoral access (78% [1015] vs 84% [1094]) in the ULM PCI group compared to the PLM PCI group (P < 0.001) (**Table 3**). Use of any kind of mechanical circulatory support (10% [134] vs 1% [17]; P <0.001) including intra-aortic balloon pump (8% [105] vs 2% [24]; P <0.001) during PCI was more common in the ULM PCI group. In the matched cohort, patients undergoing ULM PCI were more likely to undergo PCI to the left main bifurcation as compared to patients undergoing PLM PCI and a two-stent strategy was used more frequently in the ULM PCI group (22% [67/304] vs. 14% [36/250]; P = 0.003). Additionally, in the ULM PCI group more lesions (2.15 vs 1.74; P < 0.001) were treated and the use of drug eluting stents (DES) was less common (79% [1035] vs. 85% [1112]; P = < 0.001). Patients undergoing ULM PCI were less likely to undergo repeat intervention for restenosis after a prior intervention (1% [14] vs. 4% [51]; P = 0.001). Finally, in the unmatched cohort, there were significant between-group differences (P < 0.001) based on the institutional annual PCI volume in the proportion of ULM and PLM PCI performed, with low volume (<100 PCIs/year) facilities performing a minority of these cases (4% [70] vs. 7% [118]) compared to moderate volume (100-250 PCI/year) facilities (42% [663] vs. 50% [902]) and high volume (>250 PCIs/year) facilities (53% [835] vs. 43% [785]) (**Supplemental Table 3).**

**Table 3.** Procedural Characteristics for the Matched Cohort. Values are mean ± standard deviation (SD) or n (%) LM = left main; CTO = chronic total occlusion

| Procedural-Characteristics |  | Number (%) | ULM PCI<br>(n=1306) | Protected<br>LM PCI<br>(n=1306) | P Value |
| --- | --- | --- | --- | --- | --- |
| **Arterial Access site** |  |  |  |  | <0.001 |
|  | Femoral | 2109 (81 %) | 1015 (78 %) | 1094 (84 %) |  |
|  | Radial | 458 (18 %) | 273 (21 %) | 185 (14 %) |  |
|  | Brachial | 12 (0 %) | 7 (1 %) | 5 (0 %) |  |
|  | Other | 4 (0 %) | 3 (0 %) | 1 (0 %) |  |
| **Mechanical Circulatory Support (MCS)** |  |  |  |  |  |
|  | Use of any kind of MCS during CATH | 7 (0 %) | 6 (0 %) | 1 (0 %) | 0.124 |
|  | Use of intra-aortic balloon pump during CATH | 45 (2 %) | 37 (3 %) | 8 (1 %) | <0.001 |
|  | Use of any kind of MCS during PCI | 151 (6 %) | 134 (10 %) | 17 (1 %) | <0.001 |
|  | Use of intra-aortic balloon pump during PCI | 129 (5 %) | 105 (8 %) | 24 (2 %) | <0.001 |
| **Contrast volume** |  | 244 (341) | 255 (461) | 232 (125) | 0.117 |
| **Fluoroscopy time** |  | 29.5 (31.9) | 29.8 (31.1) | 29.1 (32.8) | 0.597 |
| **LM Characteristics** |  |  |  |  |  |
|  | Calcified | 999 (38 %) | 489 (37 %) | 510 (39 %) | 0.421 |
|  | Focal | 21 (1 %) | 12 (1 %) | 9 (1 %) | 0.661 |
|  | CTO | 27 (1 %) | 4 (0 %) | 23 (2 %) | 0.006 |
|  | Diffusely Diseased | 57 (2 %) | 33 (3 %) | 24 (2 %) | 0.284 |
|  | Thrombus | 36 (1 %) | 26 (2 %) | 10 (1 %) | 0.012 |
|  | Eccentric | 52 (2 %) | 34 (3 %) | 18 (1 %) | 0.036 |
|  | Serial | 10 (0 %) | 6 (0 %) | 4 (0 %) | 0.753 |
|  | Ostial | 597 (23 %) | 313 (24 %) | 284 (22 %) | 0.192 |
|  | Tortuosity | 10 (0 %) | 5 (0 %) | 5 (0 %) | 1.000 |
| **LM bifurcation** |  |  |  |  | 0.003 |
|  | 1 stent | 451 (17 %) | 237 (18 %) | 214 (16 %) |  |
|  | > 1 stent | 103 (4 %) | 67 (5 %) | 36 (3 %) |  |
| **LM Treated previously** |  |  |  |  |  |
|  | Restenosis | 65 (3 %) | 14 (1 %) | 51 (4 %) | 0.001 |
|  | Stent Thrombosis | 2 (0 %) | 1 (0 %) | 1 (0 %) | 1.000 |
| **LM lesion length, mean (SD), mm** |  | 14.5 (11.2) | 14.6 (10.6) | 14.4 (11.6) | 0.650 |
| **Number of lesions treated, mean (SD)** |  | 1.95 (1.03) | 2.15 (1.14) | 1.74 (0.871) | <0.001 |
| **Multivessel PCI** |  | 1472 (56 %) | 827 (63 %) | 645 (49 %) | <0.001 |
| **LM Intervention Characteristics** |  |  |  |  |  |
| **Balloon** |  | 2007 (77 %) | 949 (73 %) | 1058 (81 %) | <0.001 |
| **Cutting Balloon** |  | 163 (6 %) | 74 (6 %) | 89 (7 %) | 0.257 |
| **Stent- Non DES** |  | 186 (7 %) | 87 (7 %) | 99 (8 %) | 0.403 |
| **Stent- DES** |  | 2147 (82 %) | 1035 (79 %) | 1112 (85 %) | <0.001 |
| **Atherectomy** |  |  |  |  |  |
|  | Directional | 2 (0 %) | 1 (0 %) | 1 (0 %) | 1.000 |
|  | Orbital | 53 (2 %) | 35 (3 %) | 18 (1 %) | 0.026 |
|  | Rotational | 236 (9 %) | 100 (8 %) | 136 (10 %) | 0.017 |
| **Institutional Characteristics** |  |  |  |  |  |
| **Institutional annual PCI volume** |  |  |  |  | <0.001 |
|  | < 100 PCIs/year | 150 (6 %) | 63 (5 %) | 87 (7 %) |  |
|  | 100-250 PCIs/year | 1213 (46 %) | 559 (43 %) | 654 (50 %) |  |
|  | >250 PCIs/year | 1249 (48 %) | 684 (52 %) | 565 (43 %) |  |
| **On-site Cardiac Surgery back-up** |  | 2011 (77.0 %) | 1053 (80.6 %) | 958 (73.4 %) | <0.001 |

### Temporal Trends in LM PCI

Annual ULM PCI volume increased over time from 77 procedures in 2009 to 203 by 2019 at which time it comprised ∼70% of all LM PCI procedures (**Supplemental Figure S1 and Supplemental Figure S2**). Total annual LM PCI was ∼300/year throughout the study period and PLM PCI volume remained steady ranging with 149 PLM PCI in 2009 and 115 PLM PCI in 2019.

### Outcomes

Cumulative incidence of the primary endpoint (MACE at 1-year) for the matched cohorts are shown in **Table 4**. Patient undergoing ULM PCI had a higher rate of MACE (22% [289] vs. 16% [215]; P < 0.001) (**Figure 2**) and higher rate of all-cause mortality (16% [213] vs. 10% [125]; P <0.001). There were no differences in the frequency of individual outcomes such as rehospitalization for MI (4% [48] vs. 4% [48]; P = 1.000), rehospitalization for stroke (0% [4] vs. 1% [9]; P = 0.266), or revascularization (7% [94] vs. 6% [84]; P = 0.485). Patients undergoing ULM PCI vs PLM had no significant differences in peri-procedural MACE (1% [14] vs. 0% [5]; P = 0.063), and there were no significant differences in all-cause mortality (1% [13] vs. 0% [5]; P =0.098) or any of the individual MACE components **(Supplemental Table S2)**.

**Figure 2.**
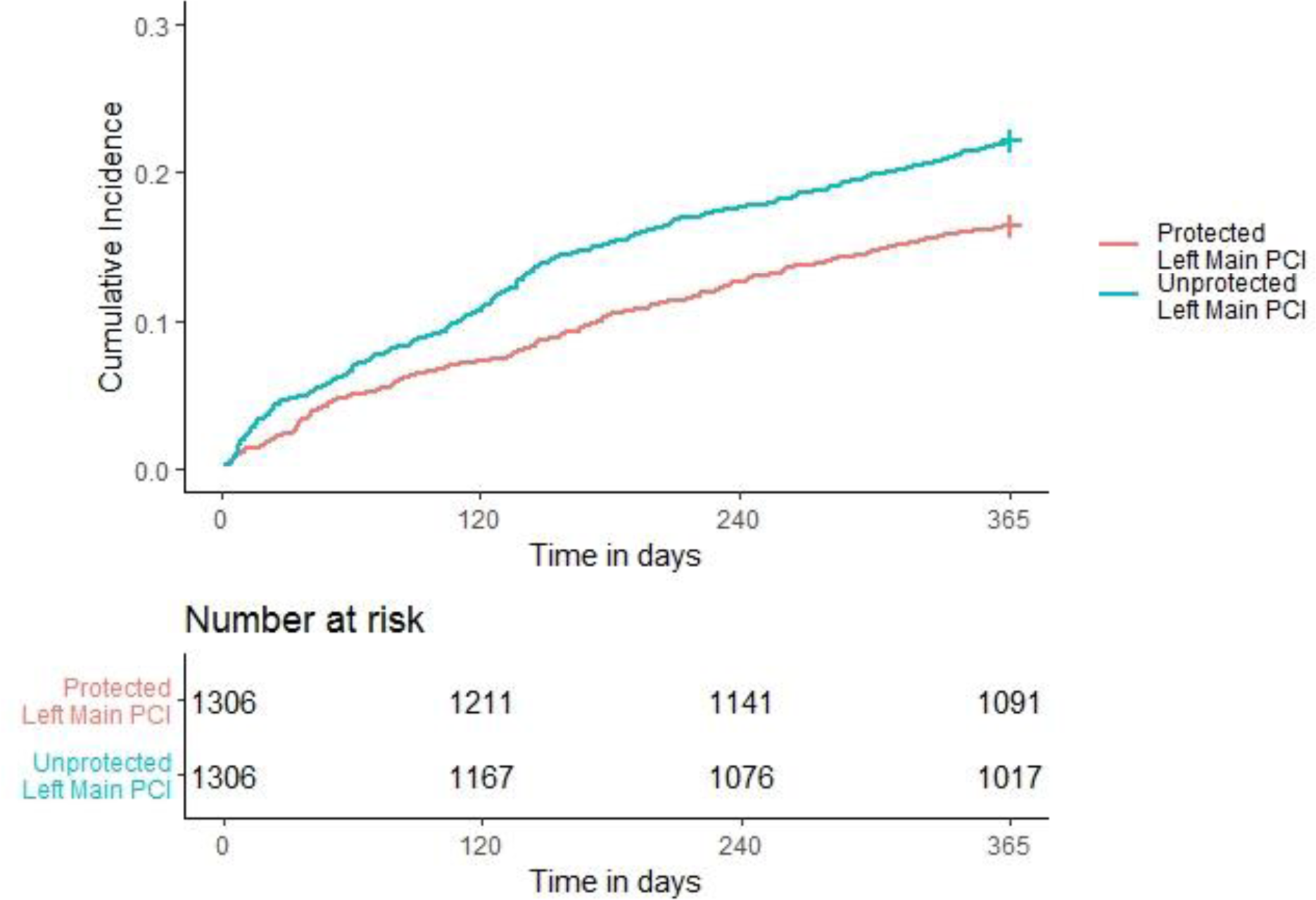
Association of PLM and ULM PCI with 1-year MACE as demonstrated by a Cumulative Incidence Plot. PCI = percutaneous coronary intervention; PLM = Protected Left Main left main; ULM = Unprotected left main.

**Table 4.** Long-term Outcomes (1-year) for Matched Cohort. Values are mean ± standard deviation (SD) or n (%) MI = myocardial infarction; ULM = unprotected left main; LM = left main

| **Outcome** | **Total** | **ULM PCI**<br>**(n=1306)** | **Protected**<br>**LM PCI**<br>**(n=1306)** | **P Value** |
| --- | --- | --- | --- | --- |
| **Primary Outcome (All-cause Mortality, rehospitalization for MI, rehospitalization for stroke or urgent revascularization)** | 504 (19 %) | 289 (22 %) | 215 (16 %) | <0.001 |
| **All-cause mortality** | 338 (13 %) | 213 (16 %) | 125 (10 %) | <0.001 |
| **Rehospitalization for MI** | 96 (4 %) | 48 (4 %) | 48 (4 %) | 1.000 |
| **Rehospitalization for stroke** | 13 (0 %) | 4 (0 %) | 9 (1 %) | 0.266 |
| **Revascularization** | 178 (7 %) | 94 (7 %) | 84 (6 %) | 0.485 |

A cumulative incidence plot was derived for 1-year MACE (**Figure 2**) and a significantly higher hazard of 1-year MACE was observed in the ULM PCI vs the PLM PCI group (HR: 1.398, (95% CI: 1.176 - 1.662); P <0.001). The approximate E-value for the estimate is 1.835 and the approximate E-value for the confidence interval is 1.484. Meaning that if there exists an unmeasured confounder that would have a hazard ratio association as large as of 1.835 with both left main PCI status and 1-year MACE, it could explain the observed association seen or an unmeasured confounder with a hazard ratio of 1.484 could move the confidence interval for ULM PCI vs PLM to include a hazard ratio of 1. Similarly, a cumulative incidence plot was derived for 1-year all-cause mortality (**Figure 3**) and a significantly higher hazard of 1-year all-cause mortality was observed in the ULM PCI vs the PLM PCI group (HR: 1.775, (95% CI: 1.482 - 1.2.207), P <0.001).

**Figure 3.**
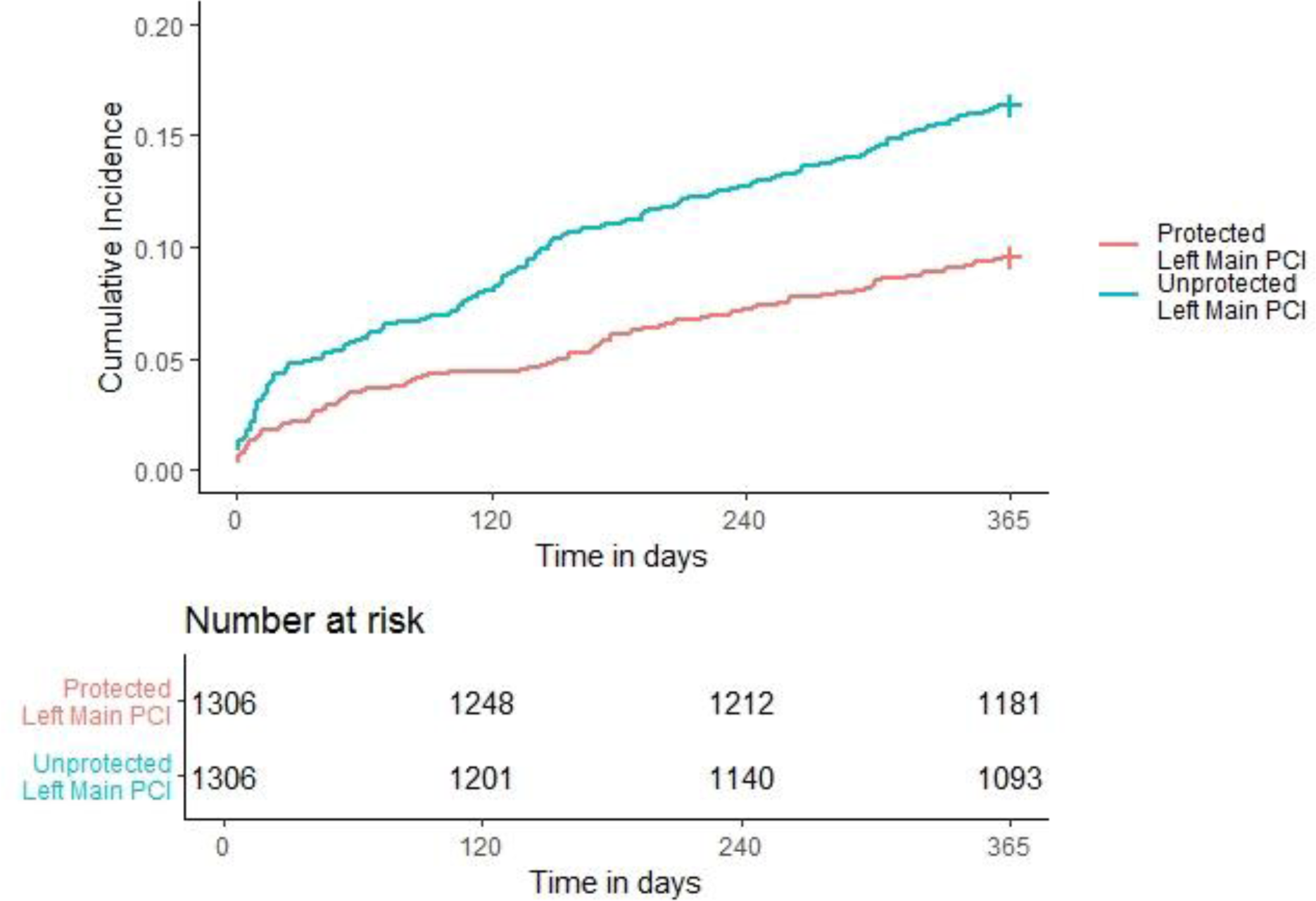
One-year All-Cause Mortality. Cumulative Incidence Plot of 1-year All-Cause Mortality for PLM and ULM PCI. PCI = percutaneous coronary intervention; PLM = Protected Left Main left main; ULM = Unprotected left main.

A cumulative incidence plot was derived for the incidence of hospitalization for MI at 12 months, adjusting for the competing risk of mortality (**Figure 4**). By the Fine-Gray hazard model, the incidence of hospitalization for MI was numerically higher for those who underwent an ULM PCI compared to those who underwent a PLM PCI, though this difference was not statistically significant (HR: 1.01, (95% CI: 0.669 – 1.50); P = 0.99). The cause-specific hazard model determined the rate of hospitalization for MI in subjects who were currently alive increased by 5% in those who underwent an ULM PCI compared to those who underwent a PLM PCI (HR: 1.05, 95% CI: 0.71 – 1.58, P = 0.35798. The approximate E-value for the estimate was 1.09 and 1.28, respectively, and the approximate E-value for the confidence interval was 1 for both models since no confounding is needed to move the confidence interval for ULM PCI vs PLM to include 1.

**Figure 4.**
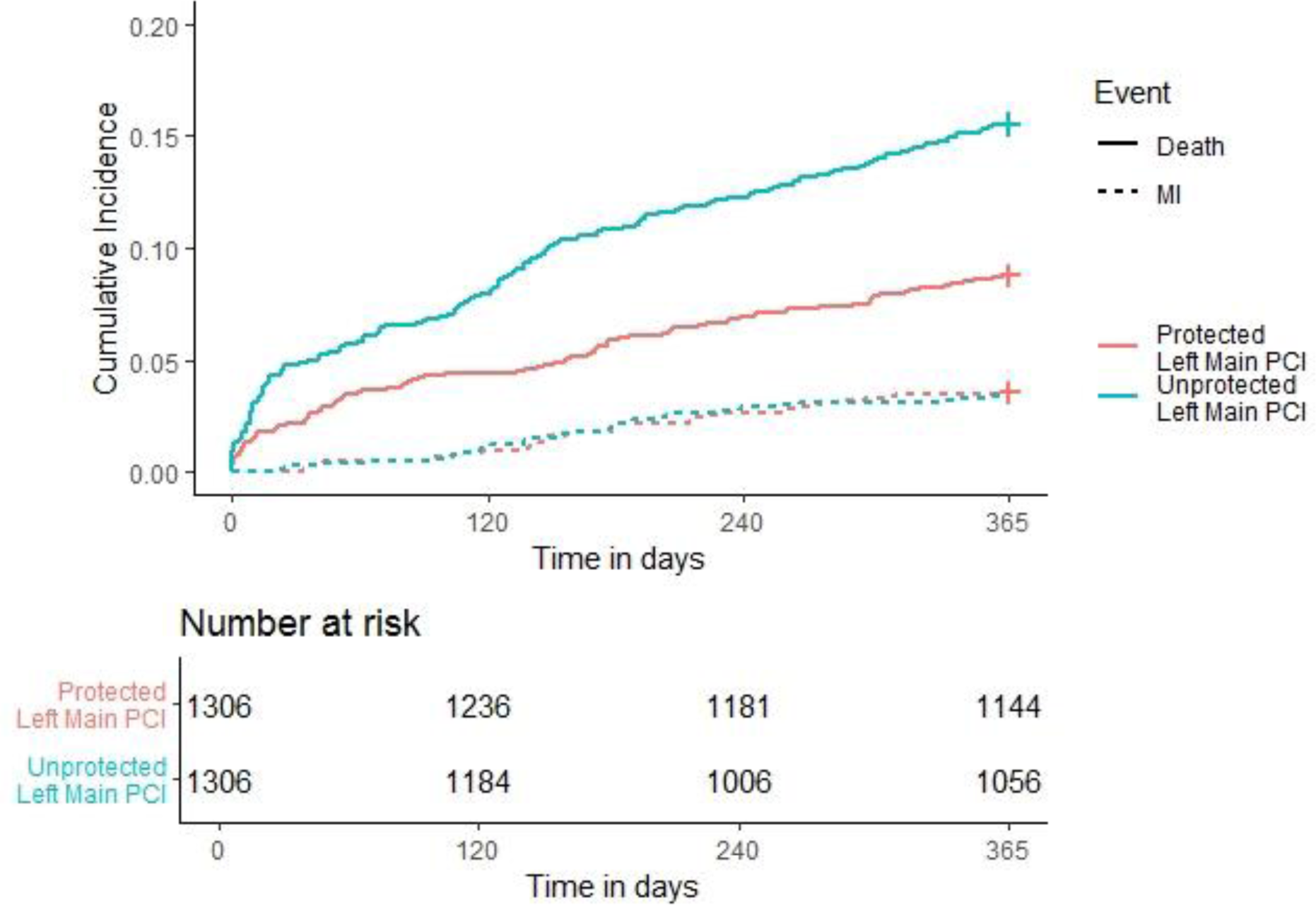
One-year Rehospitalization for MI. Cumulative Incidence Plot of 1-year Rehospitalization for MI and Competing Risk of Mortality for PLM and ULM PCI. MI = myocardial infarction; PCI = percutaneous coronary intervention; PLM = protected left main PCI; ULM = unprotected left main PCI

A cumulative incidence plot was derived for the incidence of hospitalization for revascularization at 1 year (**Supplemental Figure S3**). Using the Fine-Gray hazard model, there was no significant difference in the incidence of hospitalization for revascularization between ULM and PLM PCI (HR: 1.13, (95% CI: 0.84 – 1.51); P = 0.43). From the cause specific hazard model, the rate of hospitalization for revascularization in subjects who are currently alive increased by 18% in those who underwent an ULM PCI compared to those who underwent a PLM PCI (HR: 1.18, (95% CI: 0.88 – 1.56), P = 0.275). The approximate E-value for the estimate was 1.513 and 1.636, respectively, and the approximate E-value for the confidence interval was 1 for both models.

## Discussion

This study describes the use and outcomes of PLM and ULM PCI for LM CAD in the Veterans Health Administration. During the study period, 2,783 PLM PCI and 1,568 ULM PCI were performed, and the use of ULM PCI gradually increased over time. More than 90% of LM PCI in this cohort was performed at centers performing > 100 PCI/year. Compared to patients undergoing PLM PCI, patients in the ULM PCI cohort were older, more clinically complex, and more likely to present with ACS. After matching for baseline co-morbidities and clinical status, the primary outcome of MACE at one-year occurred more frequently in the ULM PCI group as compared to the PLM PCI group but the rate of MI or revascularization were similar. Overall, these findings suggests that there are variables contributing to all-cause mortality that are not accounted for by propensity matching. Overall, these data represent a real-world experience in a medically complex population with LM CAD, with significantly higher representation of patients with diabetes and reduced left ventricular ejection fraction compared to prior prospective registries and randomized trials.

During the 10-year study period, the total number of ULM PCI more than doubled and surpass that of PLM PCI. Overall, outcomes were better for patients undergoing PLM PCI compared to ULM PCI but there was a high incidence of MACE at one-year in both groups. This may be attributable to the baseline risk of the patient population, anatomic complexity, or a higher burden of comorbid medical conditions than prior registry-based studies. Notably, in patients undergoing PLM PCI there was a 16% incidence of MACE and 10% incidence of mortality at one year. These findings suggests that patients undergoing LM PCI after CABG may have a poor prognosis compared to patients undergoing non-LM PCI, which is in contrast to recent studies suggesting that PCI in any native vessel after CABG does not carry increased hazard.^19,20^

In this analysis our patients were more medically complex and higher risk than those enrolled in other registries of ULM PCI. Compared to a recent report of in-hospital outcomes of ULM PCI from the NCDR CathPCI registry, Veterans included in this cohort were more clinically complex with a greater proportion of heart failure, cerebrovascular disease, peripheral vascular disease, diabetes mellitus, chronic lung disease, and renal disease.^8^ Similarly, in comparison to the Drug Eluting Stent for Left Main Coronary Artery (DELTA) 2 registry, this VA cohort had a higher proportion of diabetes mellitus, lower left ventricular ejection fraction, and were more likely to present with ACS.^21^ Overall, the VA study population had a high risk for adverse cardiovascular events that could be attributed to the baseline risk profile and clinical complexity of the patients. Of note, the incidence of myocardial infarction (3% ULM PCI, 3% PLM PCI) or repeat revascularization (7% ULM PCI, 6% PLM PCI) in the current sample was similar to that of previous studies.

The recent EXCEL (Evaluation of XIENCE Versus Coronary Artery Bypass Surgery for Effectiveness of Left Main Revascularization) and NOBLE (Coronary Artery Bypass Grafting Versus Drug Eluting Stent Percutaneous Coronary angioplasty in the Treatment of Unprotected Left Main Stenosis) randomized multi-center trials compared the performance of ULM PCI to CABG. ^5,6^ In the current analysis, patients were more likely to present with ACS (55%) compared to EXCEL (38.5%) and NOBLE (17.9%) and more likely to utilize mechanical circulatory support (14.7%) as compared to EXCEL (5.6%). In our study, at one-year, the primary endpoint of MACE occurred at 24% in the ULM PCI group and 16% in the PLM PCI group. In EXCEL the primary outcome was a composite of death, stroke, or myocardial infarction and occurred at 11.5% of patients between 30 days and 3 years, and 15% of patients at 3 years; and in NOBLE the primary endpoint of major adverse cardiovascular or cerebrovascular events occurred in 7% at one year. However, VA patients undergoing ULM PCI had significantly higher mortality at one-year (16%) compared to 2.4 % in EXCEL and 2.4% in NOBLE.

How do we reconcile the difference in the outcomes of ULM PCI between this real-world multi-center experience and previous randomized clinical trials? Patients in this study had a high baseline clinical complexity and as described, a significant burden of medical co-morbidities. Two specific risk factors, diabetes mellitus and reduced left ventricular function, are known to be associated with increased mortality in patients undergoing PCI as compared with CABG, and nearly half of the current cohort is diabetic and the average left ventricular ejection fraction is reduced at 46%. This study had a high rate of transfemoral access for left main PCI (81%) versus transradial (18%), but a recent analysis from the VA healthcare system showed similar MACE and success rate between both access strategies.^22^ Despite adjusting for common co-morbidities, we found a higher frequency of MACE and all-cause mortality in the ULM PCI group compared to the PLM PCI group. These patients may have undergone PCI after being declined for surgical revascularization which could have increased the risk profile of patients included in this analysis. Data regarding surgical turndown was not available for the majority of patients. Interestingly, the incidence of myocardial infarction and stroke at one-year are similar between our analysis and other studies, raising concern that there could have been a high incidence of non-cardiovascular mortality. Finally, it is possible that the difference in outcomes could be partially attributed to differences in operator expertise with LM PCI. While in most recent randomized clinical trials higher volume operators were selected, in our study, 43-48% of PCIs were performed in high volume VA centers (>250 PCIs/year). In conclusion, this large, real-world study adds to prior registry and randomized trial data regarding LM PCI, and these data are relevant to shared decision-making with patients and multi-disciplinary cardiovascular teams regarding the role of PCI for LM CAD.

### Limitations

Registry data may be subject to errors in data entry or interpretation. VA CART data are regularly audited and incomplete or inaccurate data was omitted from this analysis. Periprocedural adverse events were captured by the VA CART Program but we were not able to assess in-hospital adverse events during the entirety of the index admission other than mortality. It is also possible that for the long-term outcomes (during the 1-year follow up period) events may have occurred outside the VA system, which would not have been captured in this analysis. There may have been additional confounders even after propensity matching that we were not able to account for, which could have contributed to the between-group differences. There may also have been selection bias with higher risk patients undergoing PCI as compared to CABG. Thus, we cannot comment on whether patients underwent PCI rather than CABG due to operator judgement, surgical turndown, or other reason. Additionally, the overwhelming majority of patients included in this study are Caucasian men and this limits generalizability to women and non-Caucasian patients. Finally, due to the balanced nature of our matched cohort, no additional variables were adjusted for in the outcome models. Therefore, interpretation of the E-values is limited to comparison of the magnitude of the treatment effect. Additionally, E-values are reported as the strength of one unmeasured confounder, though there may be multiple unknown confounders biasing the results.

### Conclusions

In the VA healthcare system patients undergoing PCI for LM CAD have a high burden of co-morbidities; however, the one-year outcomes are similar to previous real-world registries. Patients undergoing PLM PCI have better outcomes than those undergoing ULM PCI. In both groups there was a high rate of mortality and MACE at one-year despite a relatively low rate of MI or revascularization. These findings suggest that patients undergoing LM PCI in clinical practice may not reflect the patients selected for clinical trial which included patients with fewer comorbidities.

## Ethics Statement

The research reported has adhered to the relevant ethical guidelines and institutional review board approval was obtained by the Colorado Multiple Institutions Review Board and with a waiver of informed consent.

## Data Availability

The data referred to in this manuscript is availability through the Corporate Data Warehouse (CDW) from the United States Veterans Administration, and can be requested via the VA Informatics and Computing Infrastructure (VINCI) Workspace.

https://www.research.va.gov/programs/vinci/default.cfm

## Abbreviations List

LM: Left main
CAD: Coronary artery disease
PCI: Percutaneous coronary intervention
PLM: Protected left main
ULM: Unprotected left main
CABG: Coronary artery bypass graft
MACE: Major adverse cardiovascular events
VA: Veteran Affairs
CART: Clinical Assessment Reporting and Tracking
CPRS: Computerized Patient Record System
NCDR: National Cardiovascular Data Registries
MI: Myocardial infarction
BMI: Body mass index
LVEF: Left ventricular ejection fraction
NYHA: New York Heart Association
GFR: Glomerular filtration rate
STEMI: ST-segment elevation myocardial infarction
NSTEMI: non-ST-segment elevation myocardial infarction
BGA: bypass graft angiography

---

**Central Illustration:**
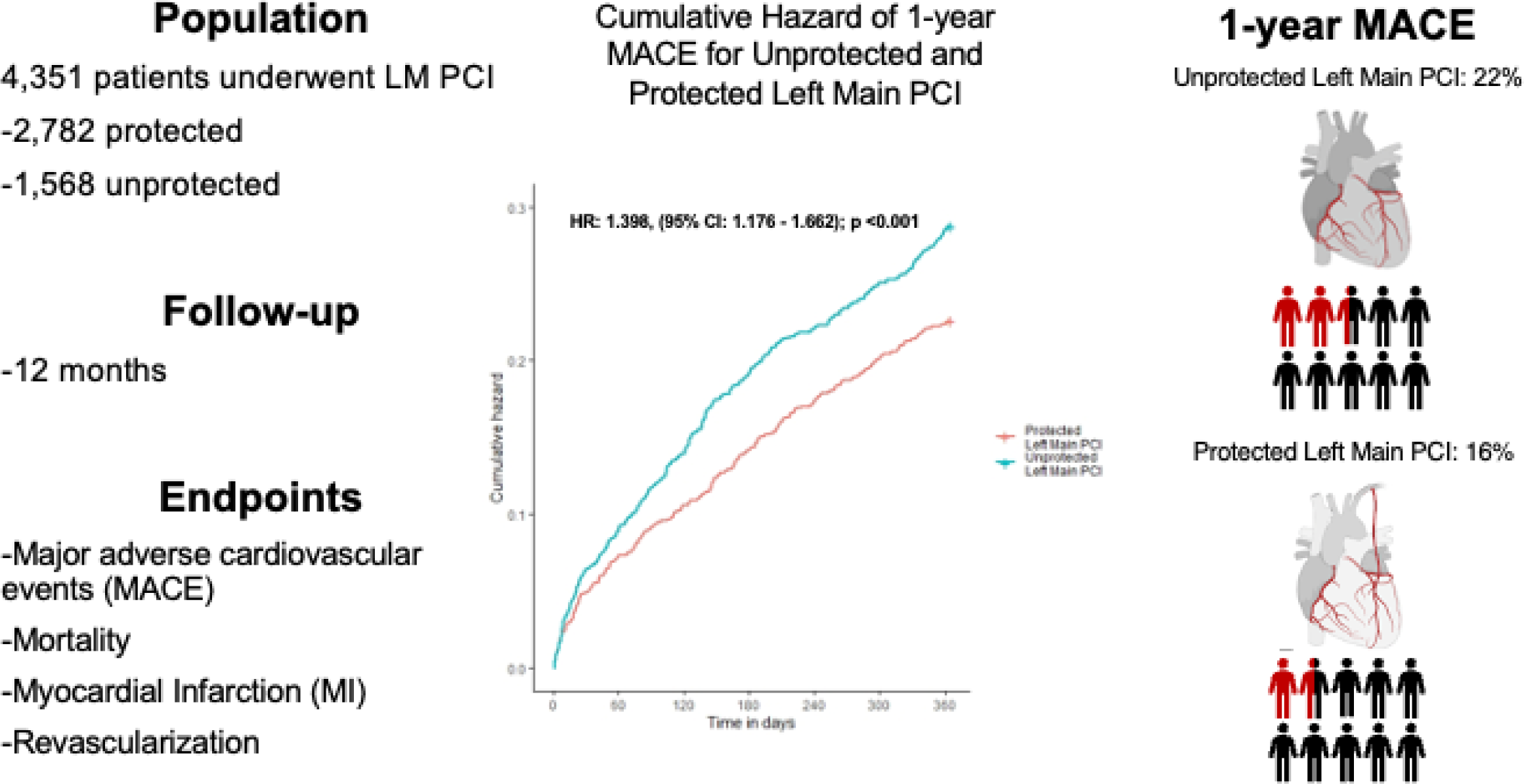
Central illustration illustrating the study design, follow-up period, and endpoints. The cumulative incidence curve demonstrates gradual diversion of the curves suggesting that factors after the initial intervention were associated with long-term adverse events. Unprotected left main PCI was associated with a 22% incidence of major cardiovascular adverse events at 1-year compared to 16% for protected left main PCI PCI = percutaneous coronary intervention; MACE = major adverse cardiovascular events; MI = myocardial infarction

## Notes

### Competing Interest Statement

The authors have declared no competing interest.

### Clinical Trial

N/A

### Author Declarations

Institutional review board approval was obtained by the Colorado Multiple Institutions Review Board and with a waiver of informed consent.

